# Stereo-encephalography-guided multi-lead deep brain stimulation for treatment-refractory obsessive compulsive disorder – study design and individualized surgical targeting approach

**DOI:** 10.1101/2025.04.17.25325961

**Authors:** Robert L. Seilheimer, Liming Qiu, Giovanna Rocchio, Young-Hoon Nho, Gustavo Campos, Andreas Horn, Camarin E. Rolle, Vivek P. Buch, T. Mindy Ganguly, Mario Cristancho, Desmond J. Oathes, Lily Brown, Bijan Pesaran, Andrew D. Krystal, Eddie F. Chang, Moses E Lee, Kai J. Miller, Nolan R. Williams, Daniel A.N. Barbosa, Katherine W. Scangos, Casey H. Halpern

## Abstract

**Introduction:** Treatment-refractory obsessive-compulsive disorder (trOCD) is a complex network disorder that may require personalized treatment strategies due to disease heterogeneity. A multi-site, multi-stage, double-blinded, randomized crossover clinical trial is underway, using stereo electroencephalography (sEEG) to guide selection of multi-nodal targets for deep brain stimulation (DBS) for trOCD.

**Objectives:** To describe the clinical trial design, emphasizing personalized surgical targeting strategies that ensure the feasibility and precision of sEEG electrode placement, and enable adequate sampling of relevant targets in trOCD for network evaluation and modulation.

**Methods:** Adults with severe trOCD (Yale-Brown Obsessive Compulsive Scale ≥ 28) who meet eligibility criteria are enrolled in this three-stage clinical trial (NCT05623306). Stage 1 involves SEEG electrode implantation in trOCD implicated regions and inpatient evaluation. Individualized probabilistic-tractography-guided target refinement is performed for surgical planning. Multimodal recordings are taken while participants stay in the psychiatric monitoring unit for 12 days. In stage 2, up to four permanent DBS electrodes are implanted followed by stimulation optimization. Stage 3 is the randomized, double blinded cross over phase.

**Expected Outcomes:** Safety, feasibility and preliminary efficacy will be assessed in this ongoing study. We anticipate that the use of sEEG to guide selection of multi-nodal targets for DBS will be safe, feasible and result in clinically meaningful improvements in symptom severity and functional impairment in trOCD.

**Discussion:** We present the clinical protocol of sEEG-guided investigation of brain networks involved in trOCD and describe our tractography-guided surgical targeting strategy designed to optimize individualized network engagement and neuromodulation.

## INTRODUCTION & STUDY RATIONALE

Obsessive-compulsive disorder (OCD) is a common psychiatric disorder with a lifetime prevalence of 2.3% in the United States and can lead to significant disability^1,2^. Despite the availability of evidence-based psychotherapy and pharmacotherapy, only an estimated 30-40% of OCD patients seek treatment, and less than half of these patients respond and few patients achieve remission with conventional interventions^1^. Of note, an accepted favorable treatment response is defined as a reduction of Yale-Brown Obsessive Compulsive Scale (Y-BOCS) of 25-35%^3^. Deep brain stimulation (DBS) targeting the anterior limb of the internal capsule (ALIC) is a Food and Drug Administration (FDA)-approved surgical intervention for treatment-refractory OCD (trOCD), under a Humanitarian Device Exemption (HDE) since 2009, but overall remains investigational^4^. Not surprisingly, outcomes of DBS in trOCD vary considerably, and the optimal stimulation target remains a subject of study amongst experts in the field^5–7^. A recent meta-analysis of 34 DBS trials in OCD comprising 352 patients with heterogenous stimulation sites (including ALIC, ventral capsule/ventral striatum, nucleus accumbens, bed nucleus of striae terminalis and subthalamic nucleus) reported an overall responder rate of 66%^8^. While this is approximately two-fold the response rate achieved with medication and psychotherapy, it is possible that optimization of stimulation and target selection could render more robust response rates and potential remissions.

OCD is now recognized as a disorder involving dysfunction across multiple, interactive brain networks^9^. While earlier models highlighted the central role of cortico-striato-thalamo-cortical (CSTC) circuitry, more recent work has implicated additional circuits – including limbic, frontoparietal, and salience networks – in contributing to the heterogeneity of OCD presentations^9,10^. Crucially, different patients may exhibit dysfunction in distinct circuits, supporting a model of OCD as a network-level disorder with individualized circuit pathology. This heterogeneity may underlie the limited success of DBS to date. Multiple circuit-level aberrancies may give rise to similar phenotypes, while shared circuit dysfunctions may produce divergent symptom profiles, underscoring the need for individualized therapies and patient-specific targeting^11,12^. To address this knowledge gap, a novel multi-site, multi-stage clinical trial is currently underway, employing stereo-electroencephalography (sEEG) to guide personalized, network-informed DBS for trOCD. This approach has recently been utilized to understand affective behaviors and personalize stimulation targets in treatment-resistant depression to personalize stimulation targets^13–15^. Here, we also describe the individualized targeting strategy used in Stage 1 of the trial, which involves inpatient sEEG monitoring to interrogate brain circuits and identify the optimal patient-specific stimulation targets.

## STUDY GOALS AND OBJECTIVES

The goal of this study is to assess the safety, feasibility, and preliminary efficacy of multi-lead DBS guided by sEEG in 10 participants suffering from severe symptoms of chronic, treatment-refractory OCD.

### Primary Objectives

- To determine each patient’s optimal targets involved in OCD symptomatology for DBS
- To assess the safety profile of a novel DBS approach, based on invasive brain monitoring mapping, compared to sham stimulation
- To assess the feasibility of a customized, novel DBS approach in brain targets previously defined by invasive brain monitoring in reducing OCD symptoms

### Secondary Objectives

- To assess the efficacy of novel stimulation parameters used in invasive brain monitoring to guide stimulation
- To assess the efficacy of a novel DBS targeting technique, based on invasive brain monitoring mapping, compared to placebo

## METHODS

This clinical trial is registered on clinicaltrials.gov (NCT05623306, NCT06347978) and first opened enrollment in April 2023. Patients with trOCD who meet eligibility criteria (Table 1) will provide informed consent to participate in this multi-stage study. The study involves an inpatient evaluation with up to 20 implanted intracranial sEEG electrodes (stage 1), followed by implantation of the permanent multi-lead DBS system and optimization of stimulation settings (stage 2). Lastly, participants enter a randomized cross-over study lasting six months to assess the efficacy of DBS therapy (stage 3) (Figure 1A-B). The trial is being conducted across three academic institutions working collaboratively: the University of Pennsylvania (Penn), Stanford University and the University of California, San Francisco (UCSF).

**Figure 1.**
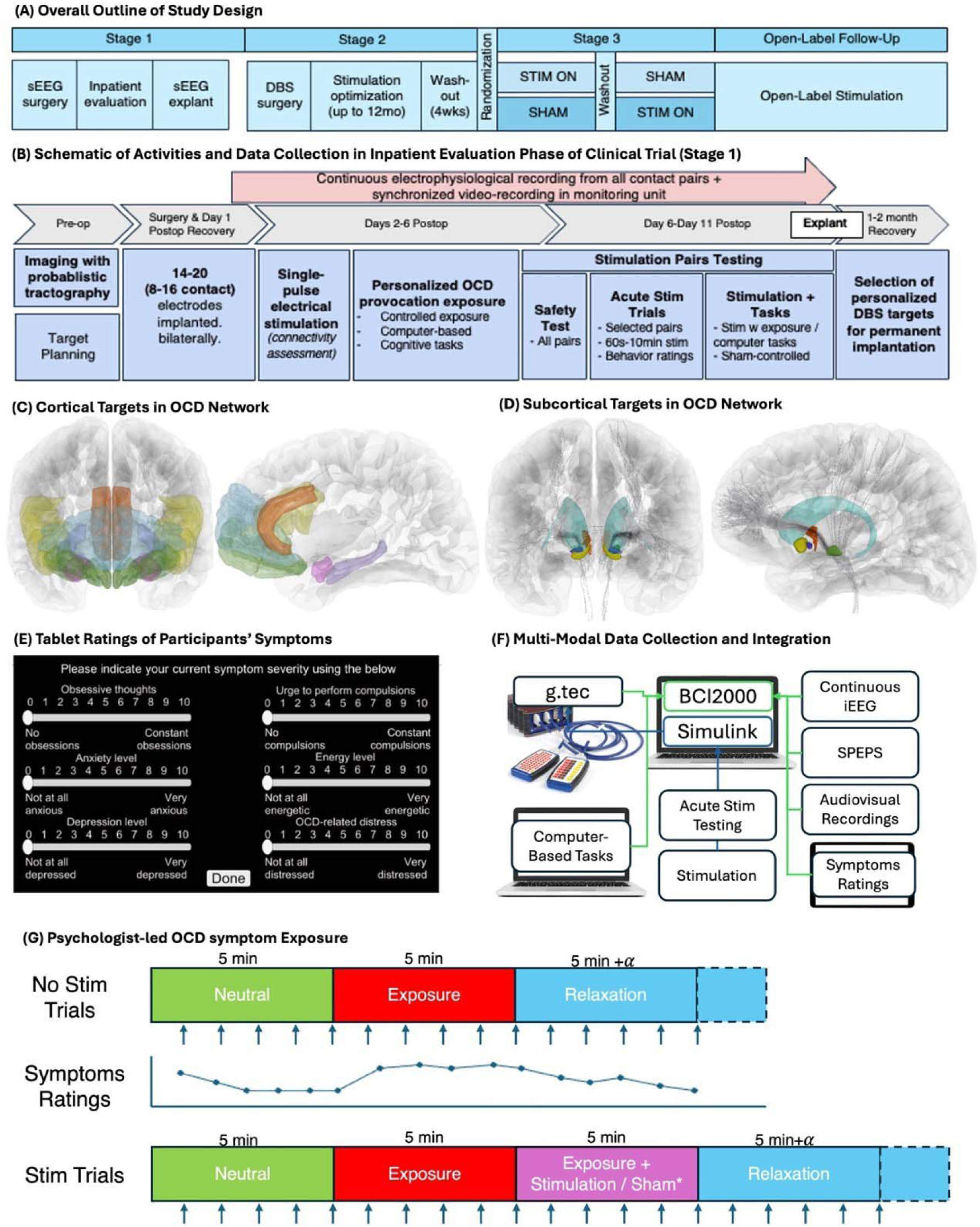
(A) Overall study outline of clinical trial of double-blinded, randomized, crossover study of stereo-encephalography guided multi-lead deep brain stimulation for treatment-refractory obsessive-compulsive disorder (NCT05623306). (B) Schematic of activities in stage 1 of clinical trial (sEEG phase). (C) Coronal (left) and mid sagittal view (right) of cortical targets in template space – orbitofrontal cortices (green), ventrolateral prefrontal cortices (yellow), frontal pole (blue), anterior cingulate cortices (orange), amygdala (pink) and hippocampi (purple). (D) Coronal (left) and mid sagittal view (right) of subcortical targets in template space – nucleus accumbens (yellow), ventral pallidum (dark blue), bed nucleus of stria terminalis (red), subthalamic nucleus (green), caudate (light blue). Streamlines of OCD response tract is overlaid in gray. (E) Screenshot of tablet interface for subject’s moment-to-moment input of multi-dimensional symptoms. (F) System set up to allow integration of multi-modal data capture in synchronized fashion. (G) A set of real-world provocation trials was designed by the psychologist, each consisting of 5-minute neutral trials and 5-minute exposure trials, followed by a relaxation/recovery period before the next round (No Stim Trials). During each trial, behavioral symptom ratings were collected every minute on a tablet (arrows). In Stim Trials, selected stimulation was applied during exposure trials to investigate its potential effect on the biomarker and symptom ratings.

**Table 1.**
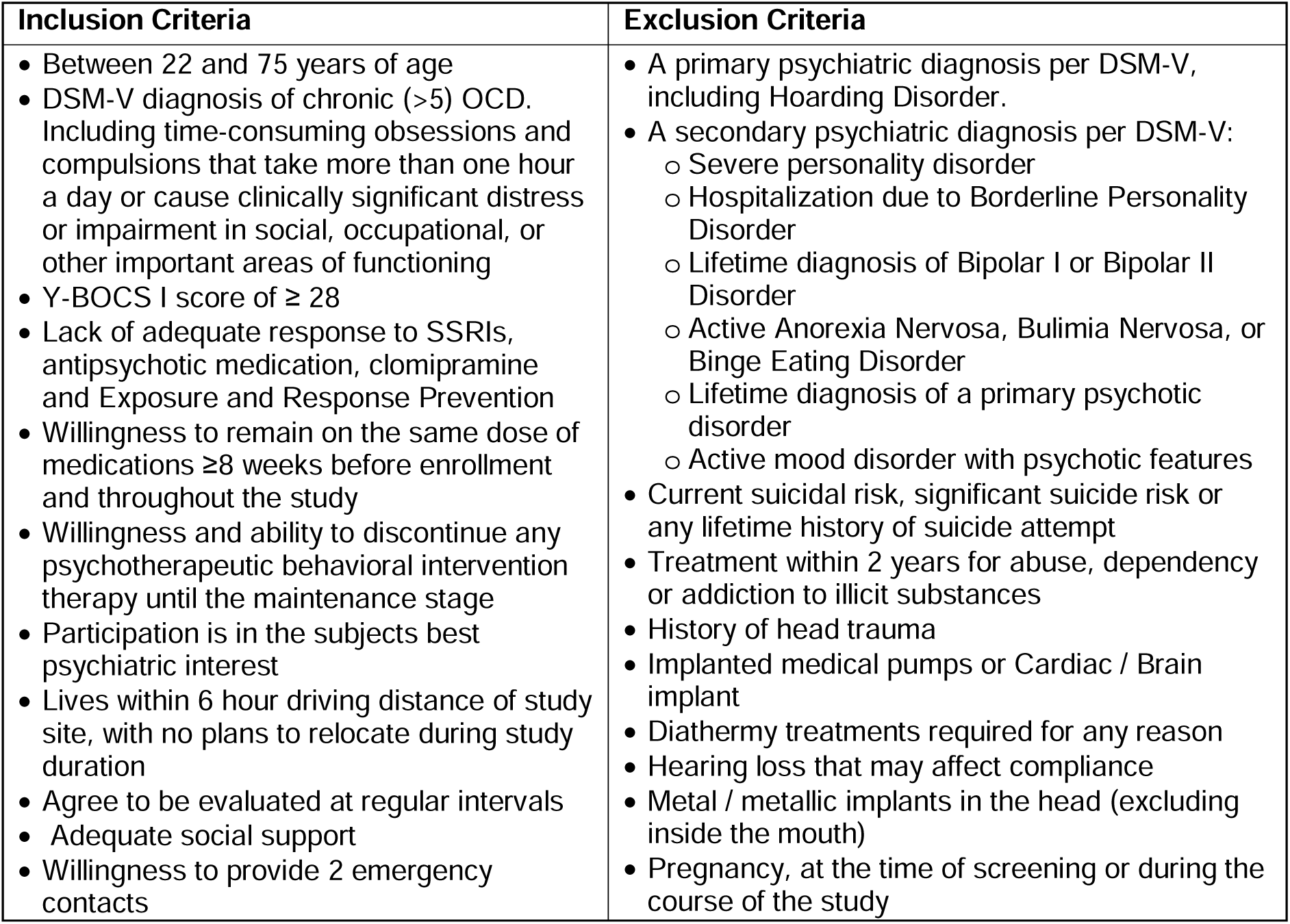
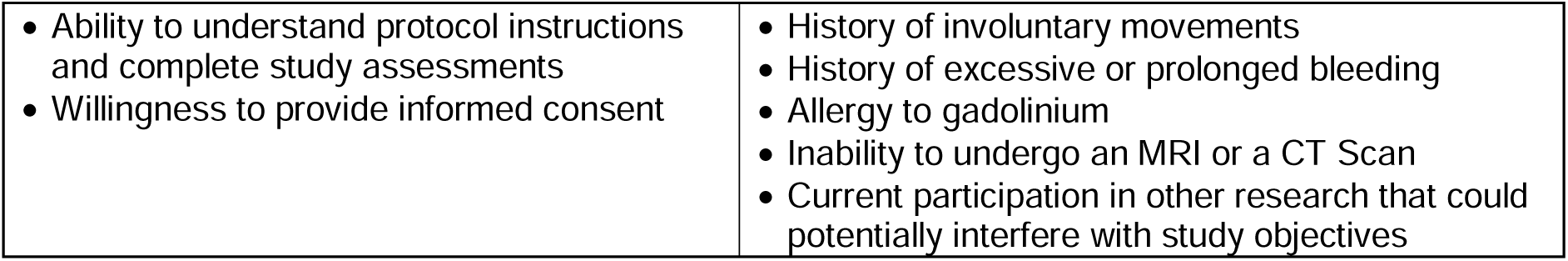
Major Inclusion and Exclusion Criteria for Subject Recruitment to Parent Clinical Trial.

### Targets Selection for sEEG lead placement

Stage 1 sEEG targets were selected through literature review as well as multi-disciplinary discussions and consensus amongst neuroanatomists, neurosurgeons, neuroimaging experts, neurologists, and interventional psychiatrists (see co-author list). Targets included structures within the fronto-striatal limbic circuitry, such as anterior and posterior orbitofrontal cortices, frontal pole, ventrolateral prefrontal cortices, anterior cingulate cortices, anterior limb of the internal capsule, anterior caudate, nucleus accumbens, ventral pallidum, subthalamic nucleus, basolateral amygdala and dorsolateral hippocampus (Figure 1C-D)^16–18^. As the aim of stage 1 evaluation is to investigate specific circuits of OCD, up to 20 sEEG electrodes are implanted to ensure sufficient coverage of cortical and subcortical regions implicated in OCD-related network dysfunction.

### Personalized Tractography-Guided Surgical Targeting

All participants will undergo high-resolution magnetic resonance imaging (MRI), consisting of T1-weighted Magnetization-Prepared Rapid Gradient-Echo (MPRAGE) with and without gadolinium contrast, T2-weighted, and Fast Gray Matter Acquisition T1 Inversion Recovery (FGATIR) sequences. Multi-shell diffusion-weighted MRI (b=1500, 3000) acquired at 1.5mm isotropic voxels and 64 diffusion directions, along with a reversed phase-encoding b0 (blip-down) image. For each individual subject, the DWI will be pre-processed using the *QSIPrep* software^19^ and co-registered to the structural T1-weighted imaging using boundary-based non-linear registration. Atlas-based anatomical segmentation will be performed following a two-step linear and non-linear co-registration using Advanced Normalization Tools (ANTs) framework with affine and asymmetric normalization^20^.

To identify critical subregions within each anatomical structure, probabilistic tractography will be performed using FMRIB Software Library (FSL)’s Probtrackx2^21^. Segmented anatomical structures will serve as seed regions and patient-specific cortical ROIs will be defined as target and waypoint masks for probabilistic tractography. Fiber orientation probability distribution will be estimated using FSL’s Bayesian Estimation of Diffusion Parameters Obtained using Sampling techniques (BEDPOSTX) model, and voxel-wise Monte Carlo sampling (5000 streamlines per voxel). The total number of streamlines reaching each ROI will be defined as the ‘waytotal’. K-means clustering will then be applied to subsegment the critical portions of the anatomical structure to guide electrode placement^22^. These resulting clusters will be converted into DICOM targeting objects using custom scripts and integrated into the Brainlab Stereotaxy Workflow (Brainlab AG, Munich, Germany)^23^. This will enable direct visualization of personalized target in native space for surgical planning (Figure 2A).

**Figure 2.**
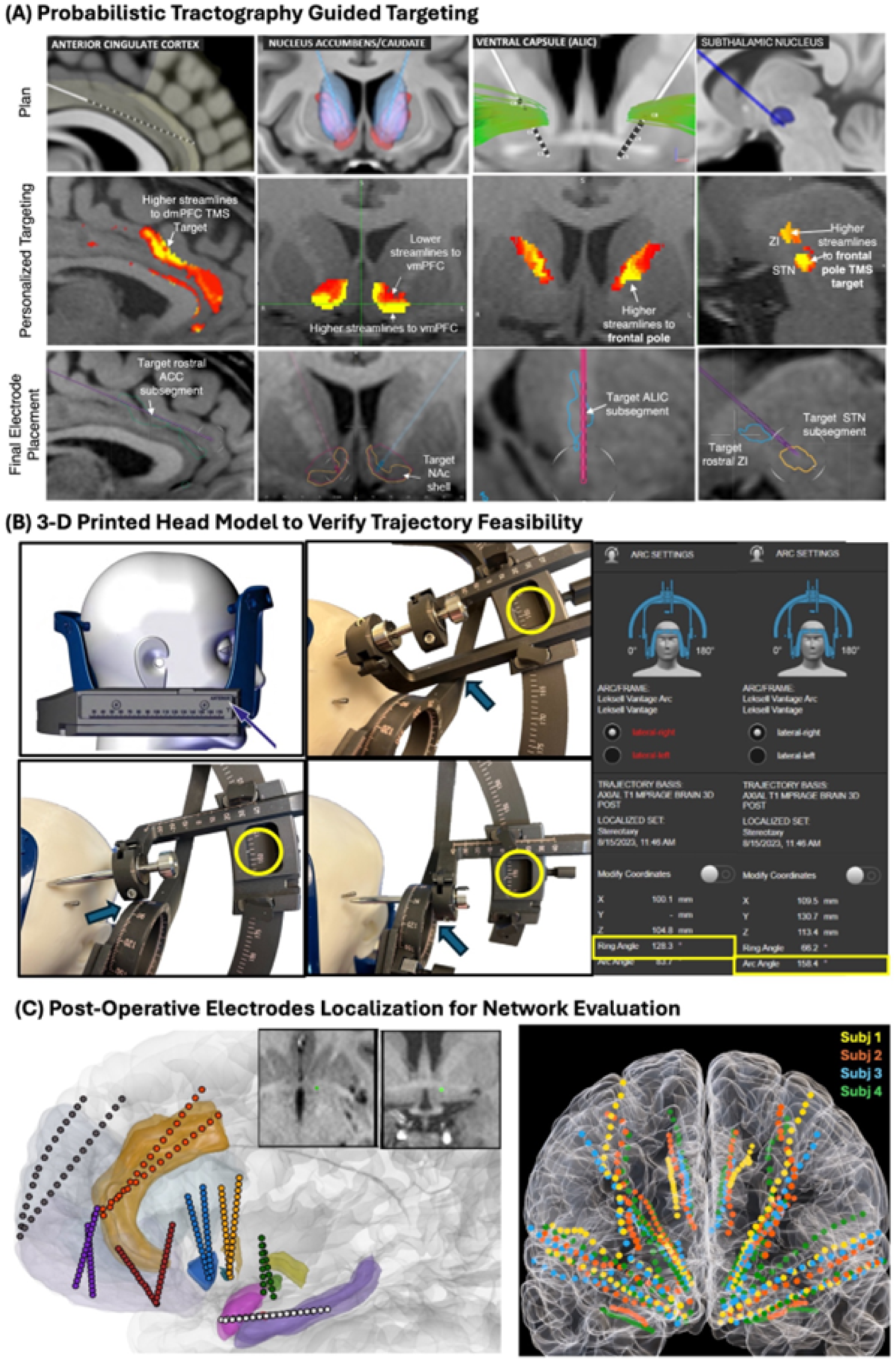
(A) Individualized probabilistic tractography was performed using high-resolution diffusion-weighted imaging, This illustrates the steps of identifying subregions with highest streamlines within four targets (anterior cingulate cortex, nucleus accumbens, ventral pallidum and subthalamic nucleus. Based on planned trajectories modelled on a standard Montreal Neurological Institute (MNI) template brain (top row), the segmented anatomical targets were used as seeds for probabilistic tractography to target frontal regions of interests. K-means clustering was used to identify subregions with the highest streamlines (yellow) within the target structure (red) (middle row). These subregions were then converted into dicom images and ported into surgical planning software (bottom row). (B) Use of subject-specific 3-dimensional head model to ensure feasibility of all trajectories and to guide frame placement. Live-size head models were 3-d printed and placed in Leksell Vantage frame for surgical rehearsal. Each trajectory was visited. Non-feasible trajectories happened when guide holders collided with stereotactic rings, or if targets exceeded frame coordinates. In such instances, trajectories were noted and revised to ensure feasibility and safety. On the day of procedure, the frame was positioned in the same manner as the model. (C) Postoperative electrodes localization in RAVE software. Left, A representative example of electrodes placement in a single subject. Inserts illustrate the electrode location of one electrode contact in axial and coronal plane. Right, Position of all electrodes from first four participants from UPenn in a standard template MNI brain.

### Surgical Implantation

Stereotactic implantation of sEEG electrodes will be performed under general anesthesia at each clinical trial site, following institutional standard protocol (i.e. frame-based technique using Leksell frame at Penn; robot-assisted with ROSA at Stanford and frame-based approach using Cosman-Roberts-Wells frame at UCSF)^13,24–26^. On the day of surgery, the participant’s head will be secured to the operative bed and stereotactic frame or robotic guidance system will be positioned. In frame-based cases, a localizer will be attached for coordinate calculation. An intraoperative computed tomography (CT) approximation will be performed using an O-arm (Medtronic, Minneapolis, MN). The CT image will then be registered to the pre-operative MRI, to obtain stereotactic coordinates of each planned trajectory. sEEG electrodes (Ad-Tech, Oak Creek, WI; PMT, Chanhassen, MN) will be implanted sequentially from posterior to anterior entry points, alternating between ipsilateral and contralateral sides. For each trajectory, a stab incision will be made with a #11 blade and a 2.4 mm drill hole made along the planned trajectory with a handheld power drill using a guide holder. An appropriately sized AdTech or PMT anchor bolt will then be placed, followed by passing an obturator to target. The sEEG electrode will then be measured to length, placed to target depth, and secured with a bolt cap. This process will be repeated for all electrodes. Two intraoperative CT will typically be performed, one after half the electrodes are placed, and one post-implantation. All electrode positions will be reviewed for accuracy of electrode position by primary site investigator, and intraoperative adjustments will be made as needed to ensure engagement of the intended anatomical targets. A radial error of less than 1.5mm from the planned trajectory will generally be considered acceptable.

### Pre-Surgical Rehearsal for the Leksell G Frame

At the time of trial initiation, the Leksell G frame was retired, and clinical experience with the newer Leksell Vantage frame was scarce. Unlike its precursor, which allowed more flexible placement (e.g. inverted or mohawk configurations), the Vantage frame imposes more restrictions, particularly for non-conventional surgical trajectories. To ensure surgical feasibility, full-scale 3-dimensional printed head model of each participant was created for pre-operative rehearsals (Figure 2B). These models allowed simulation of frame placement and identification of non-feasible trajectories. Any issues can then be addressed in advanced and rectified to avoid intraoperative complications.

### Postoperative Electrode Localization

After electrode implantation, a high-resolution, full-tissue-range head CT will be obtained. The CT will be co-registered to pre-operative MRI using BrainLAB elements as well as YAEL module of Reproducible Analysis and Visualization of iEEG software (RAVE)^27,28^. This enables calculation of precise 3D coordinates of each sEEG electrode, supporting downstream analyses (Figure 2C).

### Neuro-Psychiatric Monitoring Unit Evaluation

Following sEEG surgery, participants will be admitted to the neuro-psychiatric monitoring unit for intensive evaluation (Figure 1B). Over a 12-day inpatient stay, they will undergo a series of assessments, including single-pulse brain evoked potential connectivity assessments, psychologist-led OCD symptom provocations, acute stimulation assessments, and computer-based cognitive tasks (Table 2). Throughout the evaluation period, participants will use a tablet to provide moment-to-moment self-ratings of obsessive and compulsive symptoms, mood, energy, anxiety, and distress levels using visual analog scales (Figure 1E). All behavioral tasks are synchronized with electrophysiological recordings using digital inputs through the BCI2000 software platform^29^ and g.tec system (g.tec medical engineering GmbH, Schiedlberg, Austria) (Figure 1F). This multi-modal data collection enables detailed mapping of symptom-related brain network activity and biomarker identification across fluctuating symptom states for each participant. Following data collection, electrodes will be removed percutaneously at the bedside using aseptic techniques.

**Table 2.**
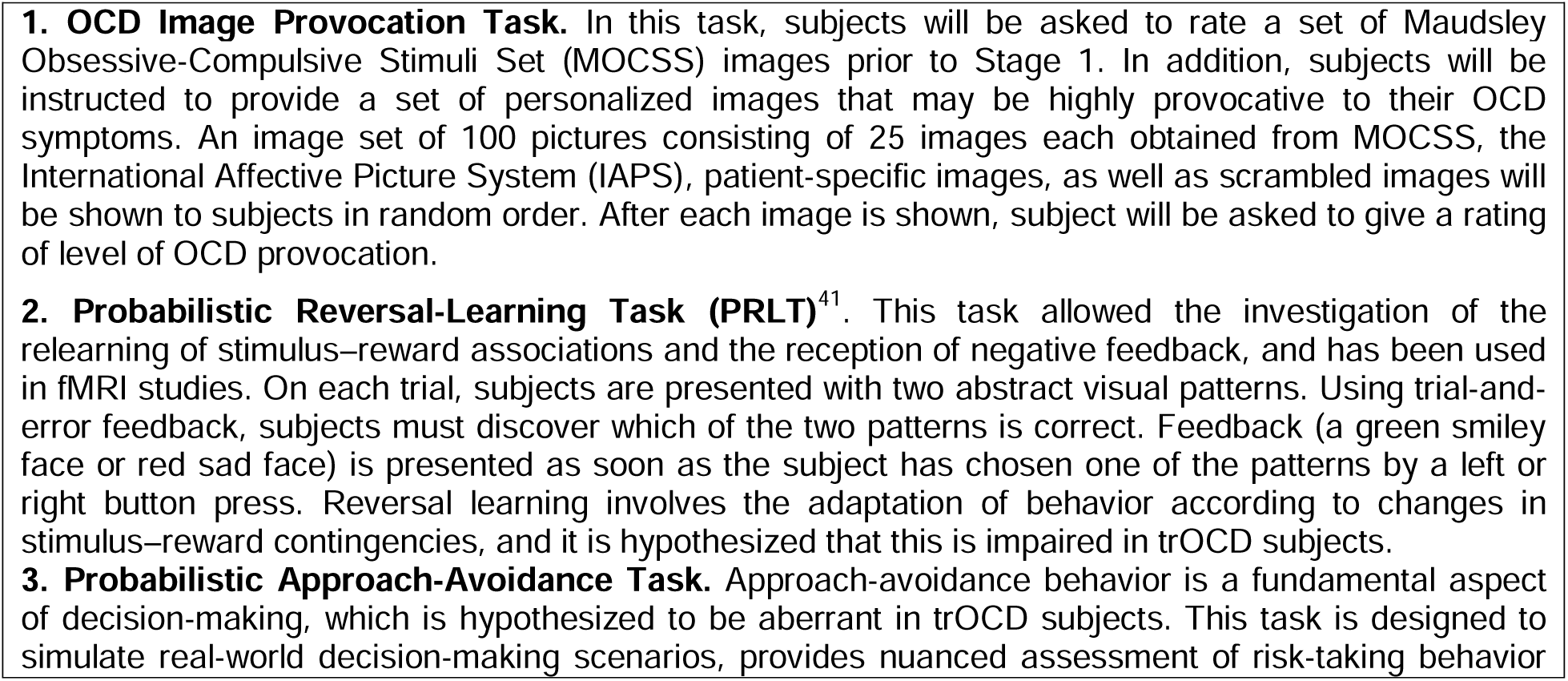
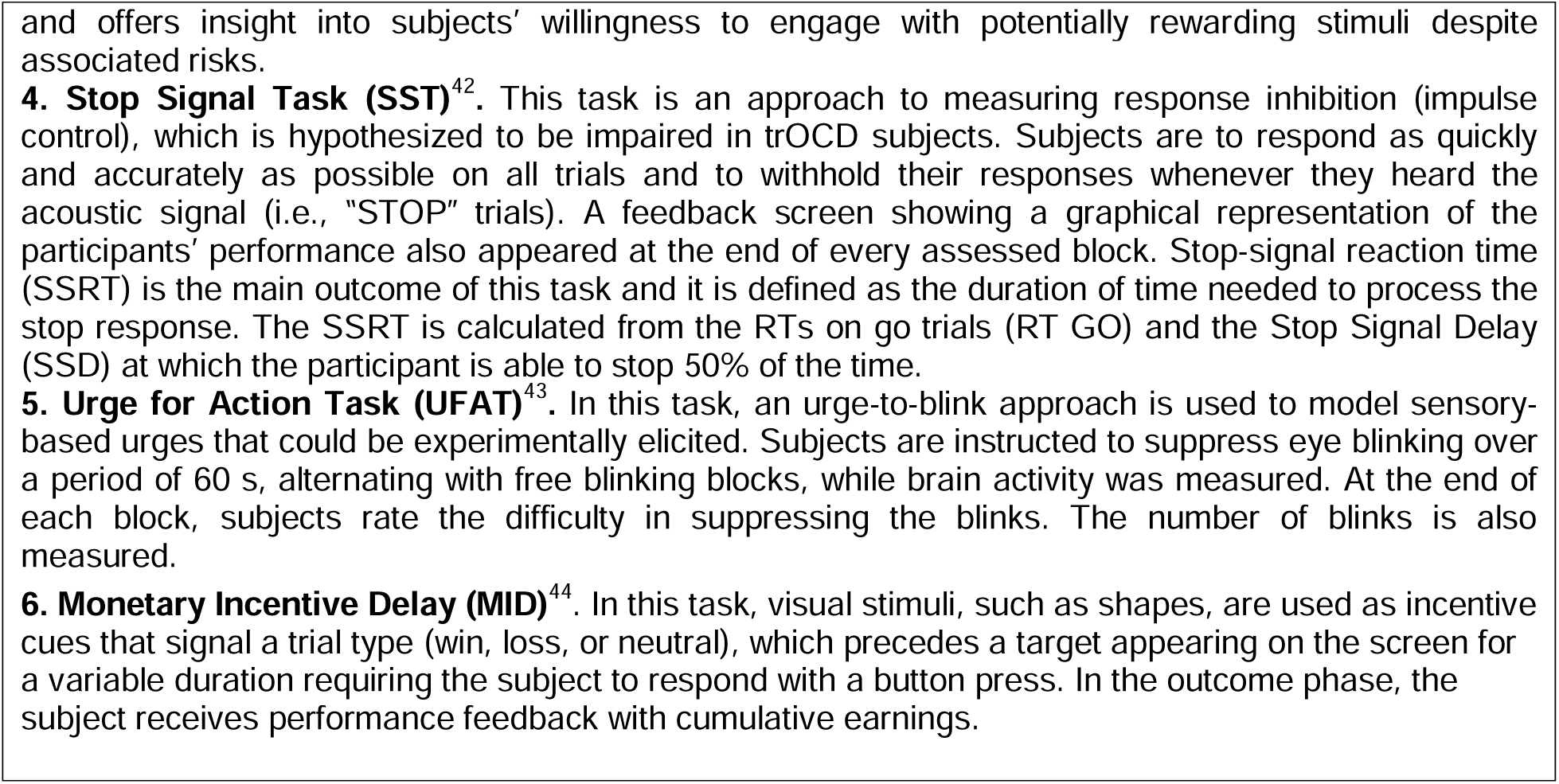
Description of Computer-based Cognitive Tasks.

#### i) Acute electrical stimulation testing

A systematic safety screening will be performed for all bipolar contact pairs using both high-frequency (100-130Hz) and low-frequency (5Hz) stimulation, at 90μs with current ranging from 0.5-6mA over 5-seconds trials. Continuous electrophysiological, video, and behavioral monitoring will be conducted during testing by a study physician and epileptologist. Electrophysiological after-discharges and any adverse clinical effects will be recorded and defined as stimulation thresholds for the remainder of the study.

Following safety validation, contact configurations will be chosen for further stimulation testing based on the relationship to targets of interests, DTI streamlines, and connectivity profiles. Selected contacts will be tested for 30-second stimulation trials and the behavioral effects will be recorded via tablet. If no adverse effects occurred, longer stimulation trials of 60-seconds, 5-minutes,10-minutes, or 20-minutes will be conducted to assess for behavioral responses. Once candidate targets are identified, randomized sham-controlled trials will be performed. Combinatorial stimulation across multiple contact pairs will also be evaluated. Both the participant and assessing physician are to be blinded to the stimulation site and condition (sham vs. active); only the study member delivering stimulation will remain unblinded.

#### ii) Psychologist-led individualized OCD Exposure

Prior to start of stage 1, a trained clinical psychologist or psychiatrist will conduct a comprehensive interview to identify each participant’s OCD symptom domains and relevant triggers. This clinician will collaborate with the study team to design personalized exposure tasks, consisting of five-minute-long neutral condition (to account for the nature of the provocation task using a neutral cue, including controlling for motor movement, speech or other factors where possible), five-minute-long symptom provocation, followed by relaxation / recovery phase. During the task, behavioral symptom ratings will be captured every minute (Figure 1G). To maximize chances of capturing symptom provoked states, individualized stimuli (videos, physical objects) will be used and repeated when highly effective or switched when less effective. To maximize discrimination between provoked and neutral/positive states, short breaks between provocations will be implemented in addition to distracting stimuli such as ‘funny/cute’ videos or conversations about hobbies, hopes for the future, etc. based on effectiveness at counteracting the symptom provocations. This paradigm serves as a platform for identifying electrophysiological biomarkers linked to OCD symptom states. We also collected symptom self-reports of spontaneous changes in OCD and related symptoms.

Stimulation effects will be further evaluated by applying selected stimulation pairs or combination stimulation during the exposure tasks. Each trial will begin with a neutral condition, followed by symptom provocation, then application of stimulation for five minutes, and finally a relaxation phase. This allows evaluation of stimulation-related behavioral modulation and concurrent neural activity.

#### iii) Brain stimulation evoked potentials (BSEPs)

On the second day after sEEG implantation (allowing for a 1-day recovery), brain stimulation evoked potentials will be performed for connectivity assessments. Single-pulse bipolar electrical stimulation (6mA, 200us) will be delivered across all adjacent contact pairs (30 trials each), and evoked responses recorded from all other electrode contacts. These evoked responses will be processed post-hoc and analyzed for measures of effective connectivity^30,31^.

After completion of stage 1 of the clinical trial, participants will undergo percutaneous removal of sEEG electrodes at the bedside per clinical practice. Following electrodes explantation, participants will enter maintenance phase, allowing for recovery. Analysis of multi-modal data obtained in Stage 1 will be performed during this time for up to six months.

### Stage 2 and 3 of Clinical Trial

Following data analysis and interpretation for each participant, the research team will recommend up to four targets for chronic DBS implantation. Standard DBS surgery will then be performed under general anesthesia using a frame-based approach previously described^24^ with either the Boston Scientific’s Vercise Genus™ R32, Medtronic Percept™ PC, or Medtronic Percept™ RC. Intraoperative awake testing will not be required, as extensive functional testing has been performed during the Stage 1 inpatient evaluation. Intraoperative brain stimulation evoked potentials may be conducted between DBS electrodes for connectivity analysis.

After a four-week post-operative recovery period, participants will return for initiation of DBS stimulation. A systematic monopolar assessment will be performed by a trained interventional psychiatrist, with DBS electrodes activated in sequential fashion. Participants will be evaluated biweekly during the DBS optimization phase (Stage 2), which lasts up to 52 weeks. The goal of the optimization phase is to achieve a Y-BOCS II score reduction of at least 35% from baseline and sustaining clinical stability for at least four weeks. Once these clinical goals are achieved, participants will advance to stage 3, the randomized cross-over phase of the study, designed to rigorously assess the clinical efficacy of DBS for OCD.

In Stage 3, DBS will first be gradually tapered off for a washout period of up to four weeks, or until the participant’s Y-BOCS-II score returns to within 20% of their pre-treatment baseline. The participants will then be randomized into either active or sham stimulation for 12 weeks. At the end of this period, a second washout phase occurs (up to four weeks), which will be followed by a crossover to the alternate condition for another 12 weeks. Throughout Stage 3, participants will undergo biweekly psychiatric evaluations. Predefined safety thresholds are set in place by the protocol to allow early rescue or progression to the next phase if needed. These include CGI-I score of 5 and active suicidal ideation with specific plan and intent as indicated with question 5 on the C-SSRS answered as ‘yes’. In the event these safety thresholds are met, the safety committee would convene to recommend appropriate actions. During the blinded stimulation phase, the optimized stimulation parameters determined from Stage 2 will be maintained. After completion of Stage 3, participants then enter an open-label follow-up phase, during which they may choose to continue DBS therapy, withdraw or explant the device. Follow-up visits will occur every eight weeks for six months.

## RESULTS AND EXPECTED OUTCOMES

### Safety and Feasibility

At time of writing, each participating site is in active recruitment. Using our deliberate, individualized surgical targeting strategy to ensure electrode placement within key network nodes, we successfully implanted all sEEG electrodes for each enrolled participant, demonstrating feasibility of the approach in Stage 1 of the clinical trial. No unexpected serious adverse events related to the surgical implantation occurred in any participant. Figure 2C depicts the normalized locations of sEEG electrode contacts for the first four subjects recruited at Penn in a common template space. All participants progressed to Stage 2 of the clinical trial with distinct DBS target combinations, reinforcing the rationale for a personalized, sEEG-guided approach.

In participants who underwent surgical implantation using the Leksell Vantage frame, the use of 3D-printed head models proved valuable for planning. These models not only helped inform optimized frame placement but enabled refinement of electrode trajectories prior to surgery. Surgical rehearsal revealed that accessing targets such as the frontal pole and orbitofrontal cortices often required the use of short posterior pins to avoid exceeding the frame’s maximum Y-coordinate (175mm). Additionally, although BrainLAB Elements software correctly reflected coordinates and arc settings, it did not fully account for potential collisions between the guide holder or guide stop with the stereotactic frame, especially with lateral-to-medial trajectories (Figure 2B, top panel). Removal of one guide holder allowed for a greater range of arc angles; however, an average collision range of 13-14 degree per side remained, rendering some lateral-to-medial trajectories infeasible (Figure 2B, bottom panels). The permissible range of ring and arc angles varied based on the specific X,Y and Z coordinates of each target^32^, underscoring the importance of feasibility checks for each trajectory.

### Treatment Efficacy

The treatment efficacy of the clinical trial will be assessed in Stage 3, using the Y-BOCS II as primary outcome measure. Secondary measures include Y-BOCS I, Obsessive-Compulsive Inventory (OCI), Montgomery-Asberg Depression Rating Scale (MADRS), Structured Hamilton-A, Visual Analogue Scales (Obsessions, Compulsions, Depression, Anxiety, Energy, and Stress), Patient Global Impression – Change Scale (PGI-C), Global Assessment of Functioning Scale (GAF), and Clinical Global Impression – Improvement Scale (CGI-I). Other clinical measures that will be reported include the following scales: the Columbia-Suicide Severity Rating Scale, the Young Mania Rating Scale, the Hamilton Depression Rating Scale, the Yale Global Tic Severity Scale, the Patient Global Impression – Severity Scale, the Clinical Global Impression – Severity Scale, the Generalized Anxiety Disorder 7-item Scale, the Mini International Neuropsychiatric Interview, and the Abnormal Involuntary Movement Scale.

## DISCUSSION

Since its initial description by Jean Talairach, and particularly over the last two decades, sEEG has become a cornerstone in the pre-surgical evaluation of epileptogenic networks^33^. As surgical safety improved, sEEG has emerged as a powerful tool for probing brain network disorders – especially those involving both cortical as well as deep structures of the brain – at high spatial (at contacts) and temporal resolution that far exceeds current non-invasive methods. Concurrently, neuropsychiatric disorders are increasingly being conceptualized as disorders of distributed brain networks. While these conditions remain incompletely understood, the application of sEEG offers a unique opportunity to interrogate network-level dysfunction from direct brain recordings. Despite its potential, sEEG has thus far been applied only in small clinical studies of depression and chronic pain conditions, where it has provided insights that informed closed-loop neurostimulation or programming strategies^13,14,31,34,35^.

Here, we present the design of the first multi-site clinical trial using sEEG to characterize brain-wide networks associated with trOCD. This trial integrates monitored, personalized OCD exposures, brain stimulation evoked potential mapping, and systematic acute stimulation testing to probe OCD-relevant networks in a rigorous, safe, and individualized manner. We emphasize the importance of precise sEEG electrode placement within targeted circuits, which is fundamental to downstream network analysis and stimulation planning. In addition, we demonstrate that a similar targeting and surgical strategy can be successfully applied across different stereotactic systems, and we provide guidance for surgical rehearsal in cases where trajectory feasibility is uncertain. The ultimate goal of the study is to synthesize multi-modal data to guide personalized, circuit-based, multi-nodal DBS therapy for trOCD. Such an approach holds promise for improving treatment outcome for this debilitating condition affecting millions of people worldwide.

Recent advances in neuroimaging and neuromodulation technologies have accelerated interests in network-informed brain stimulation therapies. Prior reports have described practical frameworks for conducting complex studies that combine imaging, physiology and behavioral data collection^26,36–38^. Our study builds upon this by incorporating individualized, tractography-informed targeting strategies designed to maximize engagement of relevant network, which is crucial to subsequent data collection and analyses using electrophysiological recordings. Using standard template atlases co-registered in individual subject’s space, we first identified specific regions of interests. These targets were then sub-segmented using probabilistic tractography to isolate subregions with the highest network connectivity. This strategy is conceptually similar to defining motor subregions within standard DBS targets for movement disorders, or the identification of subnuclei within the thalamus^39,40^. Although there is currently no standardized atlas for psychiatric DBS targeting, this individualized approach also enables incorporation of recently developed normative datasets, such as the OCD response network and connectivity sweet spots into our participants’ imaging as targeting references^6,7,17^. We did not include direct targeting of the BNST as it is a very small structure, and our consensus agreed that DBS and recordings of this structure would lack anatomical specificity. However, the volume of tissue modulated by electrodes in the VeP likely captured the BNST due to its proximity.

Our early experience with this ongoing clinical trial demonstrates the feasibility and safety of using sEEG to personalize DBS for trOCD. Data from our small initial cohort suggest that individualized targeting is not only feasible but necessary, as each participant has required a distinct combination of stimulation targets and neuromodulation protocols. Additionally, we describe a practical and reproducible frame-based targeting approach for network engagement, which could be adapted to other institutions interested in adopting similar methodologies.

## CONCLUSION

SEEG-guided exploration to identify DBS targets for neuromodulation therapy in OCD may enable a personalized understanding of the brain networks involved. In this approach, optimal placement electrode placement is essential. We present a practical targeting strategy and describe surgical planning steps that enhance confidence, improve workflow efficacy, and support safe, reproducible implantation across varying clinical settings.

## Data Availability

All data produced in the present work are contained in the manuscript.

## Acknowledgement

We gratefully acknowledge the Dr Bart Nuttin (KU Leuven), Dr Philip Mosley and Dr Terry Coyne (Queensland Brain Institute) team for their expert advice on surgical targeting. Their guidance played a key role in refining targeting strategies and ensuring the precision of electrode implantation.

## Administrative Information

Title: A Double-Blinded, Randomized, Crossover Trial of Stereoencephalography-Guided Multi-Lead Deep Brain Stimulation for Treatment-Refractory Obsessive-Compulsive Disorder

Trial Acronym: SEEG-Guided DBS for OCD

Trial Registration: NCT05623306 (registration date: 2022-10-14), NCT06347978 (registration date: 2024-03-20)

Protocol version: 12.1 (17^th^ December 2024)

Funding: Foundation for OCD Research (FFOR) and AE Foundation

Primary Sponsor: Casey H. Halpern, MD

Trial Design: Multi-stage, double-blinded randomized crossover study

Date of first enrollment: 2023-04-13

Study setting: Academic hospital

Country of recruitment: United States of America

Study population: Adult participants suffering from severe symptoms of chronic, treatment-refractory OCD

Ethic Review: Approved by the University of Pennsylvania Institutional Review Board

Estimated Completion Date: 2027-01-01

Estimated Enrollment: 10

Recruitment Status: Active recruitment

Primary Outcome(s): Change in Yale-Brown Obsessive-Compulsive Scale II (Y-BOCS II)

Secondary Outcomes(s): Y-BOCS I, Obsessive-Compulsive Inventory (OCI), Montgomery-Asberg Depression Rating Scale (MADRS), Structured Hamilton-A, Visual Analogue Scales (Obsessions, Compulsions, Depression, Anxiety, Energy, and Stress), Patient Global Impression – Change Scale (PGI-C), Global Assessment of Functioning Scale (GAF), and Clinical Global Impression – Improvement Scale (CGI-I).

## REFERENCES

1. Swierkosz-Lenart K, Dos Santos JFA, Elowe J, et al. Therapies for obsessive-compulsive disorder: Current state of the art and perspectives for approaching treatment-resistant patients. Front Psychiatry. 2023;14:1065812. doi:10.3389/fpsyt.2023.1065812

2. Stein DJ, Costa DLC, Lochner C, et al. Obsessive–compulsive disorder. Nat Rev Dis Primer. 2019;5(1):1–21. doi:10.1038/s41572-019-0102-3

3. Ramakrishnan D, Farhat LC, Vattimo EFQ, et al. An evaluation of treatment response and remission definitions in adult obsessive-compulsive disorder: A systematic review and individual-patient data meta-analysis. J Psychiatr Res. 2024;173:387–397. doi:10.1016/j.jpsychires.2024.03.044

4. Wu H, Hariz M, Visser-Vandewalle V, et al. Deep brain stimulation for refractory obsessive-compulsive disorder (OCD): emerging or established therapy? Mol Psychiatry. 2021;26(1):60–65. doi:10.1038/s41380-020-00933-x

5. Sheth SA, Mayberg HS. Deep Brain Stimulation for Obsessive-Compulsive Disorder and Depression. Annu Rev Neurosci. 2023;46:341–358. doi:10.1146/annurev-neuro-110122-110434

6. Meyer GM, Hollunder B, Li N, et al. Deep Brain Stimulation for Obsessive-Compulsive Disorder: Optimal Stimulation Sites. Biol Psychiatry. Published online December 21, 2023:S0006–3223(23)01785-7. doi:10.1016/j.biopsych.2023.12.010

7. Li N, Baldermann JC, Kibleur A, et al. A unified connectomic target for deep brain stimulation in obsessive-compulsive disorder. Nat Commun. 2020;11(1):3364. doi:10.1038/s41467-020-16734-3

8. Gadot R, Najera R, Hirani S, et al. Efficacy of deep brain stimulation for treatment-resistant obsessive-compulsive disorder: systematic review and meta-analysis. J Neurol Neurosurg Psychiatry. 2022;93(11):1166–1173. doi:10.1136/jnnp-2021-328738

9. Milad MR, Rauch SL. Obsessive-compulsive disorder: beyond segregated cortico-striatal pathways. Trends Cogn Sci. 2012;16(1):43–51. doi:10.1016/j.tics.2011.11.003

10. Shephard E, Stern ER, van den Heuvel OA, et al. Toward a neurocircuit-based taxonomy to guide treatment of obsessive-compulsive disorder. Mol Psychiatry. 2021;26(9):4583–4604. doi:10.1038/s41380-020-01007-8

11. Scangos KW, State MW, Miller AH, Baker JT, Williams LM. New and emerging approaches to treat psychiatric disorders. Nat Med. 2023;29(2):317–333. doi:10.1038/s41591-022-02197-0

12. Senova S, Clair AH, Palfi S, Yelnik J, Domenech P, Mallet L. Deep Brain Stimulation for Refractory Obsessive-Compulsive Disorder: Towards an Individualized Approach. Front Psychiatry. 2019;10. Accessed March 2, 2024. https://www.frontiersin.org/journals/psychiatry/articles/10.3389/fpsyt.2019.00905

13. Starkweather CK, Sugrue LP, Cajigas I, et al. Stereoelectroencephalography Electrode Implantation for Inpatient Workup of Treatment-Resistant Depression. Neurosurgery. 2024;95(4):941–948. doi:10.1227/neu.0000000000002942

14. Scangos KW, Makhoul GS, Sugrue LP, Chang EF, Krystal AD. State-dependent responses to intracranial brain stimulation in a patient with depression. Nat Med. 2021;27(2):229–231. doi:10.1038/s41591-020-01175-8

15. Bijanzadeh M, Khambhati AN, Desai M, et al. Decoding naturalistic affective behaviour from spectro-spatial features in multiday human iEEG. Nat Hum Behav. 2022;6(6):823–836. doi:10.1038/s41562-022-01310-0

16. Haber SN, Yendiki A, Jbabdi S. Four Deep Brain Stimulation Targets for Obsessive-Compulsive Disorder: Are They Different? Biol Psychiatry. 2021;90(10):667–677. doi:10.1016/j.biopsych.2020.06.031

17. Horn A, Li N, Meyer GM, Gadot R, Provenza NR, Sheth SA. Deep Brain Stimulation response circuits in Obsessive Compulsive Disorder. Biol Psychiatry. Published online March 20, 2025. doi:10.1016/j.biopsych.2025.03.008

18. Huey ED, Zahn R, Krueger F, et al. A Psychological and Neuroanatomical Model of Obsessive-Compulsive Disorder. J Neuropsychiatry Clin Neurosci. 2008;20(4):390–408. doi:10.1176/jnp.2008.20.4.390

19. Cieslak M, Cook PA, He X, et al. QSIPrep: an integrative platform for preprocessing and reconstructing diffusion MRI data. Nat Methods. 2021;18(7):775–778. doi:10.1038/s41592-021-01185-5

20. Avants BB, Tustison NJ, Song G, Cook PA, Klein A, Gee JC. A reproducible evaluation of ANTs similarity metric performance in brain image registration. NeuroImage. 2011;54(3):2033–2044. doi:10.1016/j.neuroimage.2010.09.025

21. Jenkinson M, Beckmann CF, Behrens TEJ, Woolrich MW, Smith SM. FSL. NeuroImage. 2012;62(2):782-790. doi:10.1016/j.neuroimage.2011.09.015

22. Barbosa DAN, Kuijper FM, Duda J, et al. Aberrant impulse control circuitry in obesity. Mol Psychiatry. Published online June 14, 2022. doi:10.1038/s41380-022-01640-5

23. Karawun: a software package for assisting evaluation of advances in multimodal imaging for neurosurgical planning and intraoperative neuronavigation | International Journal of Computer Assisted Radiology and Surgery. Accessed April 12, 2025. https://link.springer.com/article/10.1007/s11548-022-02736-7

24. Kramer DR, Halpern CH, Buonacore DL, et al. Best surgical practices: a stepwise approach to the University of Pennsylvania deep brain stimulation protocol. Neurosurg Focus. 2010;29(2):E3. doi:10.3171/2010.4.FOCUS10103

25. Ho AL, Muftuoglu Y, Pendharkar AV, et al. Robot-guided pediatric stereoelectroencephalography: single-institution experience. J Neurosurg Pediatr. 2018;22(5):1–8. doi:10.3171/2018.5.PEDS17718

26. Sheth SA, Shofty B, Allawala A, et al. Stereo-EEG-guided network modulation for psychiatric disorders: Surgical considerations. Brain Stimulat. 2023;16(6):1792–1798. doi:10.1016/j.brs.2023.07.057

27. Magnotti JF, Wang Z, Beauchamp MS. RAVE: Comprehensive open-source software for reproducible analysis and visualization of intracranial EEG data. NeuroImage. 2020;223:117341. doi:10.1016/j.neuroimage.2020.117341

28. Wang Z, Magnotti JF, Zhang X, Beauchamp MS. YAEL: Your Advanced Electrode Localizer. eNeuro. 2023;10(10):ENEURO.0328-23.2023. doi:10.1523/ENEURO.0328-23.2023

29. Schalk G, McFarland DJ, Hinterberger T, Birbaumer N, Wolpaw JR. BCI2000: A General-Purpose Brain-Computer Interface (BCI) System. IEEE Trans Biomed Eng. 2004;51(6):1034-1043. doi:10.1109/TBME.2004.827072

30. Miller KJ, Müller KR, Valencia GO, et al. Canonical Response Parameterization: Quantifying the structure of responses to single-pulse intracranial electrical brain stimulation. PLoS Comput Biol. 2023;19(5):e1011105. doi:10.1371/journal.pcbi.1011105

31. Scangos KW, Khambhati AN, Daly PM, et al. Closed-loop neuromodulation in an individual with treatment-resistant depression. Nat Med. 2021;27(10):1696–1700. doi:10.1038/s41591-021-01480-w

32. Krüger MT, Terrapon APR, Hoyningen A, Kim CHO, Lauber A, Bozinov O. Posterior Fossa Approaches Using the Leksell Vantage Frame with a Virtual Planning Approach in a Series of 10 Patients-Feasibility, Accuracy, and Pitfalls. Brain Sci. 2022;12(12):1608. doi:10.3390/brainsci12121608

33. Dasgupta D, Miserocchi A, McEvoy AW, Duncan JS. Previous, current, and future stereotactic EEG techniques for localising epileptic foci. Expert Rev Med Devices. 2022;19(7):571–580. doi:10.1080/17434440.2022.2114830

34. Sheth SA, Bijanki KR, Metzger B, et al. Deep Brain Stimulation for Depression Informed by Intracranial Recordings. Biol Psychiatry. 2022;92(3):246–251. doi:10.1016/j.biopsych.2021.11.007

35. Zhao H, Zhang S, Wang Y, et al. A pilot study on a patient with refractory headache: Personalized deep brain stimulation through stereoelectroencephalography. iScience. 2024;27(2):108847. doi:10.1016/j.isci.2024.108847

36. Noecker AM, Mlakar J, Bijanki KR, et al. Stereo-EEG-guided network modulation for psychiatric disorders: Interactive holographic planning. Brain Stimulat. 2023;16(6):1799–1805. doi:10.1016/j.brs.2023.11.003

37. Noecker AM, Mlakar J, Petersen MV, Griswold MA, McIntyre CC. Holographic visualization for stereotactic neurosurgery research. Brain Stimulat. 2023;16(2):411–414. doi:10.1016/j.brs.2023.02.001

38. Allawala AB, Bijanki KR, Adkinson J, et al. Stereo-Electroencephalography–Guided Network Neuromodulation for Psychiatric Disorders: The Neurophysiology Monitoring Unit. Oper Neurosurg. 2024;27(3):329. doi:10.1227/ons.0000000000001122

39. Ewert S, Plettig P, Li N, et al. Toward defining deep brain stimulation targets in MNI space: A subcortical atlas based on multimodal MRI, histology and structural connectivity. NeuroImage. 2018;170:271–282. doi:10.1016/j.neuroimage.2017.05.015

40. Akram H, Dayal V, Mahlknecht P, et al. Connectivity derived thalamic segmentation in deep brain stimulation for tremor. NeuroImage Clin. 2018;18:130–142. doi:10.1016/j.nicl.2018.01.008

41. Cools R, Clark L, Owen AM, Robbins TW. Defining the Neural Mechanisms of Probabilistic Reversal Learning Using Event-Related Functional Magnetic Resonance Imaging. J Neurosci. 2002;22(11):4563–4567. doi:10.1523/JNEUROSCI.22-11-04563.2002

42. Martoni RM, Risso G, Giuliani M, et al. Evaluating Proactive Strategy in Patients With OCD During Stop Signal Task. J Int Neuropsychol Soc. 2018;24(7):703–714. doi:10.1017/S1355617718000267

43. Er S, C B, M L, et al. The buildup of an urge in obsessive-compulsive disorder: Behavioral and neuroimaging correlates. Hum Brain Mapp. 2020;41(6). doi:10.1002/hbm.24898

44. Knutson B, Adams CM, Fong GW, et al. Anticipation of Increasing Monetary Reward Selectively Recruits Nucleus Accumbens. J Neurosci. 2001;21(16):RC159-RC159. doi:10.1523/JNEUROSCI.21-16-J0002.2001

